# How does treatment coverage and proportion never treated influence the success of *Schistosoma mansoni* elimination as a public health problem by 2030?

**DOI:** 10.1101/2023.10.20.23297322

**Authors:** Klodeta Kura, Nyamai Mutono, Maria-Gloria Basáñez, Luc E. Coffeng, S.M. Thumbi, Roy M. Anderson

**Affiliations:** London Centre for Neglected Tropical Disease Research, London, UK; Department of Infectious Disease Epidemiology, School of Public Health, Faculty of Medicine, St Mary’s Campus, Imperial College London, UK; MRC Centre for Global Infectious Disease Analysis, School of Public Health, Imperial College London, London, UK; Centre for Epidemiological Modelling and Analysis, University of Nairobi, Nairobi, Kenya; Paul G Allen School for Global Health, Washington State University, Pullman, USA; Department of Public Health, Erasmus MC, University Medical Center Rotterdam, Rotterdam, The Netherlands; Institute of Immunology and Infection Research, University of Edinburgh, Edinburgh, UK

**Author notes:** Co-first authors (Klodeta Kura, Nyamai Mutono).

**Keywords:** schistosomiasis, mass drug administration, MDA, modelling, elimination as a public health problem, community-wide, never treatment

## Abstract

**Background:** The 2030 target for schistosomiasis is elimination as a public health problem (EPHP), achieved when the prevalence of heavy intensity infection among school-aged children (SAC) reduces to <1%. To achieve this, the new World Health Organization (WHO) guidelines recommend a broader target of population to include pre-school (pre-SAC) and adults. However, the probability of achieving EPHP should be expected to depend on patterns in repeated uptake of mass drug administration (MDA) by individuals.

**Methods:** We employed two individual-based stochastic models to evaluate the impact of school-based and community-wide treatment and calculated the number of rounds required to achieve EPHP for *Schistosoma. mansoni* by considering various levels of the population never treated (NT). We also considered two age intensity profiles, corresponding to a low and high burden of infection in adults.

**Results:** The number of rounds needed to achieve this target depends on the baseline prevalence and the coverage used. For low and moderate transmission areas, EPHP can be achieved within seven years if NT ≤10% and NT <5%, respectively. In high transmission areas, community wide treatment with NT<1% is required to achieve EPHP.

**Conclusions:** The higher the intensity of transmission, and the lower the treatment coverage, the lower the acceptable value of NT becomes. Using more efficacious treatment regimens would permit NT values to be marginally higher. A balance between target treatment coverage and NT values may be an adequate treatment strategy depending on the epidemiological setting, but striving to increase coverage and/or minimise NT can shorten programme duration.

## Introduction

Schistosomiasis is a neglected tropical disease (NTD) caused by the trematode worm *Schistosoma* and transmitted through dermal contact with water contaminated by cercariae, the infectious stage of schistosomes, which are released by the intermediate host snail [1]. The major disease-causing species are *S. mansoni, S. haematobium* and *S. japonicum*. In 2016, schistosomiasis was estimated to account for 1.9 million disability adjusted life years, likely a gross underestimate [2,3]. In 2021, the World Health Organization (WHO) Roadmap on NTDs proposed elimination of schistosomiasis as a public health problem (EPHP; defined as prevalence of heavy intensity infection reducing to <1% in SAC) in all 78 endemic countries by 2030 [4].

Globally, 240 million people reside in areas endemic for schistosomiasis, with 91% of the population at risk living in Africa [5]. Efforts to control and eliminate the disease have been predominantly through preventive chemotherapy (PC) treatment with praziquantel (PZQ), which kills the adult worms [4]. Over the years, PZQ has been targeted at school aged children (SAC, 5-14 years) in endemic settings, who have the highest risk of infection [6]. To achieve EPHP, the 2022 WHO treatment guidelines recommend inclusion of adults, pre-school aged children (pre-SAC) and women of reproductive age (including pregnant women from the first trimester), with a target of at least 75% treatment coverage of eligible population per treatment round [7]. The treatment of pre-SAC would require a new, paediatric formulation of PZQ. The proportion of population never treated (NT) after continuous rounds has been reported to influence the success of mass drug administration (MDA) campaigns, and the likelihood of achieving elimination targets for helminthiases [8].

Mathematical models have been used to estimate the impact of MDA in achieving disease elimination, while accounting for the pre-control endemicity, treatment coverage and frequency [9,10]. However, the implications of the proportion NT are understudied.

In this work, we used mathematical models to provide insights into the impact of NT on achieving the 2030 EPHP target. Specifically, we assessed what proportion of NT would influence the likelihood that schistosomiasis programmes achieve EPHP (defined as achieving <1% heavy-intensity prevalence in SAC) target, different treatment regimens (annual, bi-annual), intensity profile and coverage levels.

## Methods

We used two individual-based stochastic transmission models developed by Imperial College London (ICL) [11–13] and the University of Oxford (SCHISTOX) [14] to simulate the effect of different levels of NT and MDA coverage among SAC and community on the probability of reaching EPHP for low (<10%), moderate (10-50%) and high baseline prevalence (>50%) areas as defined by the magnitude of the basic reproduction number, R_0_ (ranging from 1.2 to 4). Both models had similar processes, except for one important difference. The ICL model assumed that the number of eggs produced is a non-linear function (density-dependent egg production) of the female worm burden assuming monogamous sexual reproduction. In contrast, SCHISTOX assumed that the number of eggs produced is proportional to the number of worm pairs (male and female worms). Both models were calibrated with the same baseline settings, by varying the R_0_ in the ICL model, and the overall contact rate (one term in the denominator of R_0_) in the SCHISTOX model.

We modelled a population of 500 individuals without migration, and various levels of NT (measured after five rounds of MDA) among eligible individuals (ranging from 0% to 40%), depending on treatment coverage, following Dyson et al [8].

We assessed the impact of coverage for 60% and 75% of the community (treating those aged ≥2) and 75% of SAC (5-14 years), with annual treatment frequency in low to moderate prevalence areas and biannual (6-monthly) in high prevalence areas. We also considered two age-intensity profiles of infection, corresponding to low or high burden of infection in adults [10,15,16]. Table 1 provides parameter values used in the models.

**Table 1.** Parameter values for *Schistosoma. mansoni*.

| Parameter | SCHISTOX | ICL | Reference |
| --- | --- | --- | --- |
| Fecundity (eggs/female/sample) | 0.34 | 0.34 | [12,17,18] |
| Aggregation parameter | 0.04-0.24 | 0.04-0.24 | [6,16] |
| Density dependent fecundity | 0.0007 | 0.0007 | [6,19] |
| Worm life span (years) | 5.7 | 5.7 | [6,12,20] |
| Low adult burden setting:<br>Age specific contact rates for 0-5,<br>5-10, 10-16, 16+ years old | 0.01, 1.2, 1, 0.02 | 0.01, 1.2, 1, 0.02 | [19,21] |
| High adult burden setting:<br>Age specific contact rates for<br>0-5, 5-12, 12-20, 20+ years old | 0.01, 0.61, 1, 0.12 | 0.01, 0.61, 1, 0.12 | [19,21] |
| Drug efficacy | 86.3% | 86.3% | [22] |
| Contact rate | 0.03-0.18 | - | - |
| Basic reproduction number | - | 1.2-4 | - |
| Population size | 500 | 500 | - |

At the end of the 20-year treatment duration, we evaluated the heavy intensity infection to determine whether the proposed EPHP threshold had been met. Each scenario was run 500 times and we considered EPHP to be achieved when 90% of the simulations were below 1% of heavy intensity prevalence in SAC, which was measured by single Kato-Katz on two samples per individual, regardless of the burden of infection in adults.

## Results

In low prevalence areas, treating 60% of the community with 1% NT would achieve the EPHP target within five years, regardless of the burden of infection in adults. Increasing the coverage to 75% of the community increases the probability of elimination (EPHP) and reduces the required number of rounds to achieve the target by one year (Table 2). To achieve EPHP within seven years, the NT should not exceed 15% in low adult burden and 10% in high adult burden settings when treating the community (those aged ≥2 years). Achieving the same target while treating 75% of SAC only would require the NT to be 15% and 1% in settings with low and high burden of infection in adults, respectively.

**Table 2.**
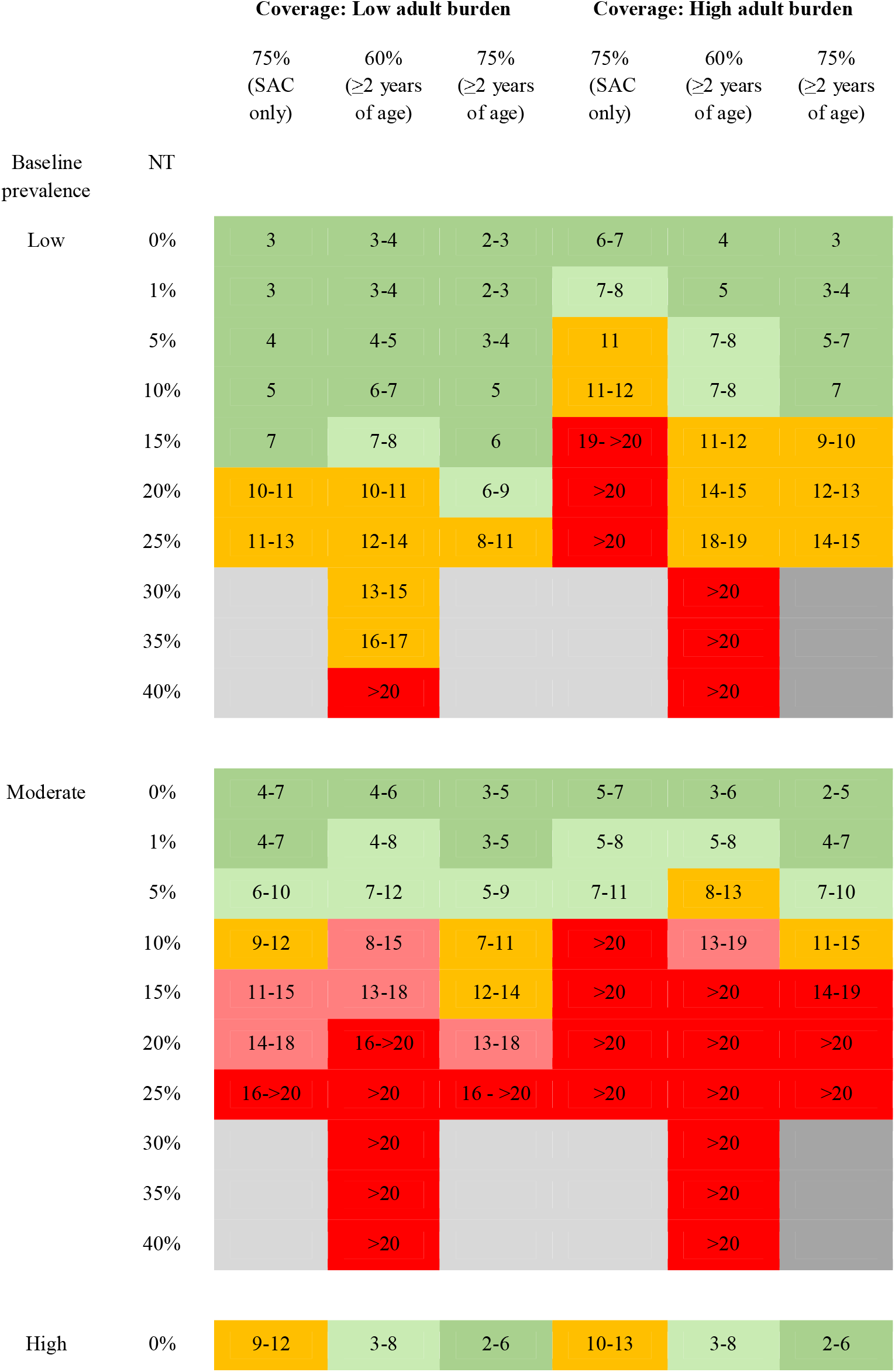
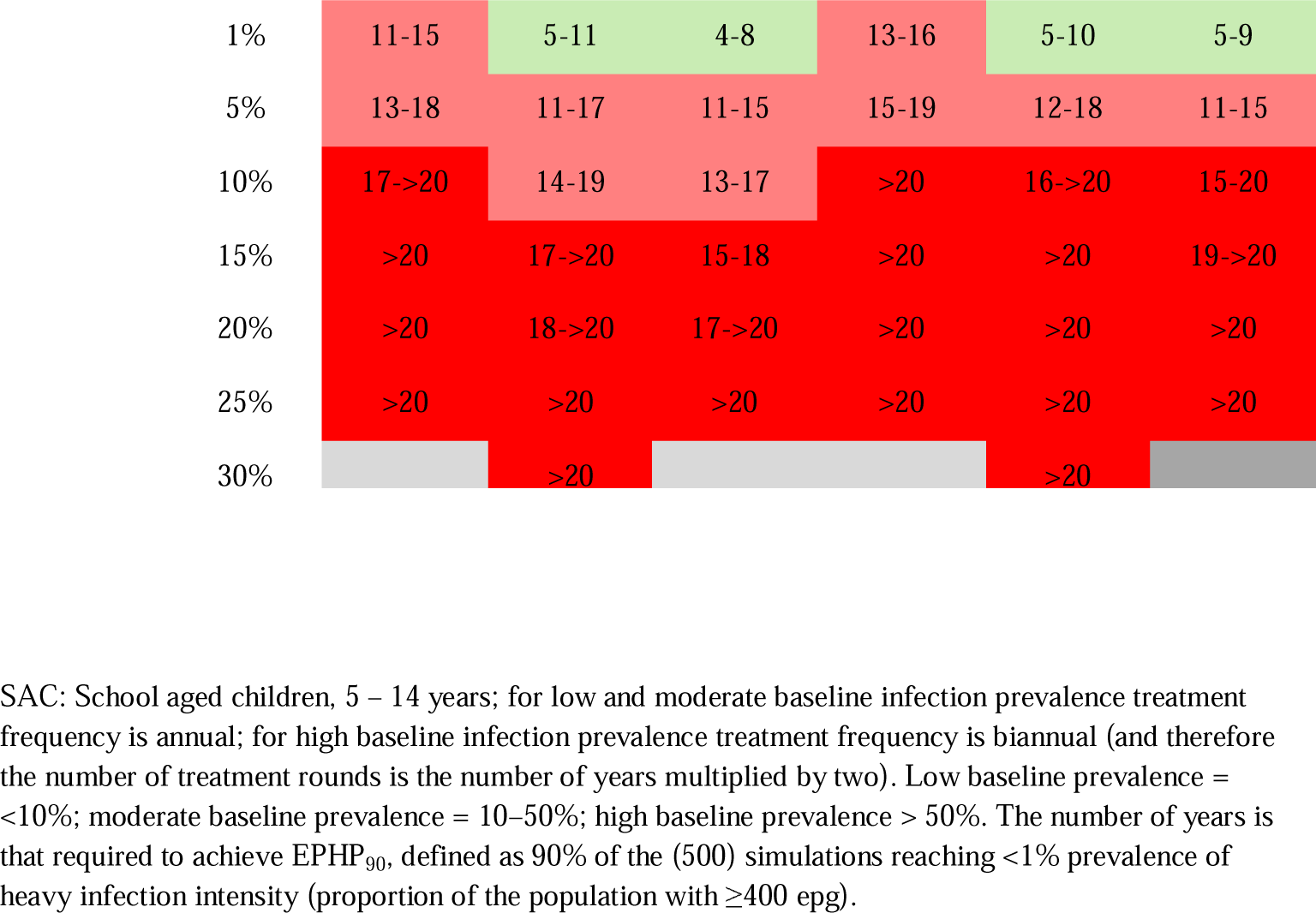
Model recommended treatment strategies for achieving elimination as a public health problem (EPHP) for low and high burden of infection in adults with different proportions of population never treated (NT). Coverage and NT values are among eligible population. Results are generated using the ICL and SCHISTOX models. The dark green shade shows EPHP achieved within seven years, orange within eight to 14 years, and red above 14 years. The light green and light red shades show the borderline values within seven and 8-14 years and 8-14 years and >14 years respectively. The grey areas show scenarios that cannot be simulated, based on the treatment coverage.

In moderate prevalence areas, the EPHP target would be achieved within seven years for all treatment strategies with NT=0% (random treatment), regardless of the burden of infection in adults. Increasing the NT to 1% increases the required number of treatment rounds to achieve the target by one year, whereas for NT = 10% more than ten years would be required (Table 2).

In high prevalence areas, the EPHP target would be achieved within seven years by treating 75% of the community (those aged ≥2 years), regardless of the burden of infection in adults provided that NT=0%. Treating SAC only in high prevalence areas would not achieve EPHP target within seven years, and a proportion NT = 1% would require more than 12 years of biannual treatment to achieve EPHP.

For a low burden of infection in adults, the success of a SAC only treatment programme depends on the baseline prevalence, and the NT proportion. For baseline prevalence above a threshold (67% for ICL and 76% for SCHISTOX), an increase in SAC coverage and inclusion of adults is recommended to achieve the target within seven years of treatment. Specific results from each model are presented in Supplementary Material Table S1. Elimination probability (EPHP) results for a high prevalence setting when NT=0% are shown in Figure S1.

## Discussion

We find that community-wide treatment including the use of the new formulation of praziquantel to treat pre-SAC can achieve elimination as a public health problem within a short time frame provided MDA coverage is good and individual compliance to treatment is effectively random at each round. Independent of MDA coverage, the outcome depends on the burden of infection in adults and the baseline prevalence (determined by the magnitude of R_0_). The higher the MDA coverage and treatment compliance, (Table 2) the lower the number of rounds required to achieve this target.

Despite the target being achieved in some areas for different treatment strategies, there is a high risk of resurgence following MDA cessation if control efforts are not maintained. The worm aggregation in a community is unevenly distributed, and it is challenging to measure the variability after MDA treatment. The worm aggregation may increase after many rounds of MDA if there is a small proportion of people with heavy-intensity infection that has never been treated. These individuals are a reservoir of infection and increase the risk of resurgence. To prevent resurgence, it is important to maintain EPHP with reduced efforts (less frequent or lower coverage of MDA) or move toward the interruption of transmission goal [10, 21]. The likelihood of maintaining the EPHP target will critically depend on the strategy adopted and the transmission setting, whereby more intense efforts are required in high transmission areas.

For a given NT value, treatment coverage is an important driver of programme duration: the greater the coverage of eligible population the shorter the projected number of years to achieve EPHP. This is because as prevalence falls in the majority of the population, infection levels in NT individuals also decrease due to a lower incidence of new infections through lower transmission, and natural death of existing worms that are replenished at a lower rate. As long as there are only a few NT individuals harbouring reproductively active worms, transmission in the overall population may fall sufficiently low that eventually, infection levels in NT individuals are not able to sustain infection for the entire population above 1% prevalence of heavy intensity infection in SAC.

There is a clear need for more studies of individual compliance patterns in PZQ MDA-treated communities, as very few longitudinal studies of compliance have been conducted [23]. In future work we will use data from the ongoing Geshiyaro project in Ethiopia which is following a large population treated with PZQ over many rounds of MDA and recording individual adherence behaviours [24].

Whilst our models consider closed populations, human movement between communities (either as short-term commuting or long-term migration, including population displacement as a result of civil unrest and/or climate change) can hamper the success of MDA programmes by reducing the probability of elimination (or increase the rate of resurgence upon cessation of MDA) due to spatial diffusion between communities with differing levels of treatment coverage [22]. This is particularly important when programmes transition from EPHP towards elimination of transmission. It is also important to consider the sensitivity of the diagnostic technique. In this study, the prevalence of infection was measured by Kato-Katz which has a low sensitivity in detecting infection at very low prevalence areas. Alternative diagnostic techniques such as the point-of-care circulating cathodic antigen (POC-CCA) could be helpful as it has a greater sensitivity at low prevalence than Kato-Katz [25–27].

Additional interventions, such as improving water, sanitation, and hygiene (WASH), the future use of an efficacious vaccine (if one were to become available) and/or snail control could reduce the number of years of MDA required to achieve EPHP.

## Data Availability

All data produced in the present work are contained in the manuscript

## Authors’ contributions

K.K.: conceptualization, formal analysis, investigation, methodology, visualization, project administration, writing—original draft, writing—review and editing; N.M.: conceptualization, formal analysis, investigation, methodology, visualization, writing— original draft, writing—review and editing; M.G.B: conceptualization, writing—review and editing; L.E.C: conceptualization, methodology, writing—original draft, writing—review and editing; S.M.T., R.M.A.: conceptualization, methodology, visualization, supervision, writing—review and editing.

## Abbreviations

epg: eggs per gram of faeces
EPHP: elimination as a public health problem
EPHP_90_: probability of reaching EPHP in 90% of model runs
EOT: elimination of transmission
MDA: mass drug administration
NT: proportion (of eligibles) never treated
NTD: neglected tropical disease
pre-SAC: pre-school age children
PZQ: praziquantel
*R*_0_: basic reproduction number
SAC: school-age children
WHO: World Health Organization

